# Clinical and Genetic Evaluation of Suicide Death with and without Interpersonal Trauma Exposure

**DOI:** 10.64898/2026.04.14.26350901

**Authors:** Eric T. Monson, Andrey A. Shabalin, Emily DiBlasi, Michael J. Staley, Erin A. Kaufman, Anna R. Docherty, Amanda V. Bakian, Hilary Coon, Brooks R. Keeshin

## Abstract

**Importance:** Suicide is a leading cause of death in the United States with risk strongly influenced by Interpersonal trauma, contributing to treatment resistance and clinical complexity.

**Objective:** To assess clinical and genetic factors in individuals who died from suicide, with and without interpersonal trauma exposure.

**Design:** Individuals who died from suicide with and without trauma were compared in a retrospective case-case design. Prevalence of 19 broad clinical categories was assessed between groups. Results directed selection of 42 clinical subcategories, and 40 polygenic scores (PGS) for further assessment. Multivariable logistic regression models, adjusted for critical covariates and multiple tests, were formulated. Models were also stratified by age group (<26yo and ≥26yo), sex, and age/sex.

**Setting:** A population-based evaluation of comorbidity and polygenic scoring in two suicide death subgroups.

**Participants:** A total of 8 738 Utah Suicide Mortality Research Study individuals (23.9% female, average age = 42.6 yo) who died from suicide were evaluated, divided into trauma (*N* = 1 091) and non-trauma exposed (*N* = 7 647) individuals. A subset of unrelated European genotyped individuals was also assessed in PGS analyses (Trauma *N* = 491; Non-trauma *N* = 3 233).

**Exposures:** “Trauma” is here defined as interpersonal trauma exposure, including abuse, assault, and neglect from International Classification of Disease coding.

**Main Outcomes and Measures:** Prevalence of comorbid clinical sub/categories and PGS enrichment in trauma versus non-trauma exposed suicide deaths.

**Results:** Overall, trauma-exposed individuals died from suicide earlier (mean age of 38.1 yo versus 43.3 yo; *P* <0.0001) and were disproportionately female (38% versus 21%, OR = 3.3, CI = 2.9-3.8). Prevalence of asphyxiation and overdose methods, prior suicidality, psychiatric diagnoses, and substance use (OR range = 1.3-3.7) were elevated in trauma exposed individuals who died from suicide. Genetic PGS were also elevated in trauma-exposed individuals who died from suicide for depression, bipolar disorder, cannabis use, PTSD, insomnia, and schizophrenia (OR range = 1.1-1.4) with ADHD and opioid use showing uniquely elevated PGS in trauma exposed males (OR range = 1.2-1.4).

**Conclusions and Relevance:** Results demonstrated multiple convergent lines of age- and sex-specific evidence differentiating trauma-exposed from non-trauma exposed suicide death. Such findings suggest unique biological backgrounds and may refine identification and treatment of this high-risk group.

## Introduction

Suicide claims in excess of 700,000 lives annually throughout the world^1^ and is the second leading cause of death among adolescents and young adults, aged 10-34, within the United States.^2^ Despite efforts to improve detection and treatment of mental illness and reduce stigma and barriers to treatment, suicide rates have risen over the past several decades.^3^ Prediction of risk of suicide death, an inherently preventable outcome, remains little better than chance.^4^

One of the greatest challenges in suicide death prediction stems from the complexity and heterogeneity of suicidality. The definition of suicidality as represented within the literature includes suicidal ideation, attempt, and death.^5^ Many researchers focus their studies on more common outcomes like suicidal ideation and attempts. However, over half of suicide deaths occur without prior clear suicidal behavior and 54% of individuals who die from suicide have no known psychiatric diagnoses.^6^ Additionally, >90% of individuals who attempt suicide do not die from suicide.^7^ Thus, examining cases of suicide death can provide unique insight into important risk factors, including genetic vulnerabilities and environmental and clinical precipitants.^8^

Risk for suicidality can be conceptualized within a diathesis – stress model.^9^ The diathesis, or underlying biological risk for suicide, is thought to be polygenic in nature, arising from many sites of genetic variation for which we have evolving evidence.^8,10,11^ This biological diathesis is hypothesized to mediate specific behavioral phenotypes, including response patterns to stress and tendencies toward impulsiveness and/or aggression.^12-14^ The stress component may arise through any number of perceived environmental stressors experienced throughout life, including remote, chronic, and acute/triggering stressors and exposures. Complex interaction of genetic and environmental suicidality risk factors contributes to the complexity of the current literature and the limited overlap of results.

Prior interpersonal trauma exposure is a potent etiological suicide risk factor that impacts specific populations. Previous studies have demonstrated lasting negative outcomes stemming from interpersonal trauma including overall worsened physical and psychiatric disease trajectories,^15^ greater resistance to traditional treatments,^16,17^ and increased risk for suicidal behavior.^18-20^ Age and sex are also hypothesized to be significant factors in predicting outcomes for patients exposed to interpersonal trauma. For example, “complex” variations of PTSD,^21^ are suspected to result from chronic/repeated traumatic exposures that begin in childhood and are identified more frequently in females. These features may indicate that specific groups have an underlying biological liability for dysregulated responses to environmental stress.^22^

In sum, the complexity of suicidality makes prediction of death difficult, but evaluation of suicide death coupled with consideration of specific exposures may reduce complexity and allow more targeted prevention, intervention, and management strategies. Exposure to interpersonal trauma results in a clinically distinct group with substantially increased risk for suicide.^20^ In addition, the diathesis-stress model has never been studied within suicide death through incorporation of both genetic and exposure/clinical data. In this study, we utilize unique suicide death clinical and genetic data resources to investigate how suicide death in individuals with a history of interpersonal trauma differ from other individuals who die from suicide.

## Materials and Methods

### Sample Selection

The Utah Suicide Mortality Research Study (USMRS) is a unique data resource derived from >12 500 population ascertained individual who died from suicide.^10,20,23^ 8 738 individuals with linked electronic medical records were selected for this study. 3 714 unrelated European individuals had genotype data available. This study was approved by the Utah Resource for Genetic and Epidemiological Research Committee at the University of Utah and Institutional Review Boards from the University of Utah, Intermountain Health, and the Utah Department of Health and Human Services.

### Phenotype definition and demographic analyses

#### Individuals were divided into two groups

**Trauma-Exposed Suicide** “TES” (*N* = 1 091) and **Non-Trauma-Exposed Suicide** “NTES” (*N* = 7 647). The TES group was defined via International Classification of Diseases (ICD) versions 9 and 10 coding that documented prior trauma through a diagnosis of post-traumatic stress disorder, acute stress disorder, experienced abuse, neglect, or assault (see Supplemental Table S1 for ICD codes). The NTES group was composed of all other individuals. Demographics analysis utilized t-tests for age and number of diagnostic codes per individual, chi-square tests for sex, and logistic regression, accounting for age and sex, for cause of death (dependent variable; obtained from death certificate data). Groups were also divided into two age groups and their demographics compared: mature adults (aged ≥ 26yo; range: 26-97yo; TES *N* = 874, NTES *N* = 6 193) and young adults and adolescents (aged <26yo; range: 7-25.9yo; TES *N* = 217, NTES *N* = 1 454).

### Clinical Analyses

TES and NTES were compared for prevalence of diagnoses within 19 primary clinical categories representing broad physiologic and pathologic processes (see Supplemental Table S2), including one female-specific category: pregnancy and related diagnoses. Prevalence comparisons were conducted by formulating a series of logistic regression models with the diagnostic category treated as the binary dependent variable, and TES/NTES classification as the independent variable. Prevalence comparisons were also performed within 42 clinical category subsets enriched for TES in the primary analysis. See Supplemental Table S3 for clinical subgroup diagnostic codes. Secondary analyses were performed within all clinical categories stratified by age, <26yo and ≥26yo ^24^ (two models), sex (two models), and age and sex (four models). All models were adjusted for sex (excepting sex-stratified models), age, and number of diagnostic codes as each of these factors differed significantly between groups (see Table 1). All tests were corrected for multiple testing via the Benjamini-Hochberg method with a false discovery rate (FDR) of 0.10. All statistical analyses were performed within R.^25^

**Table 1:**
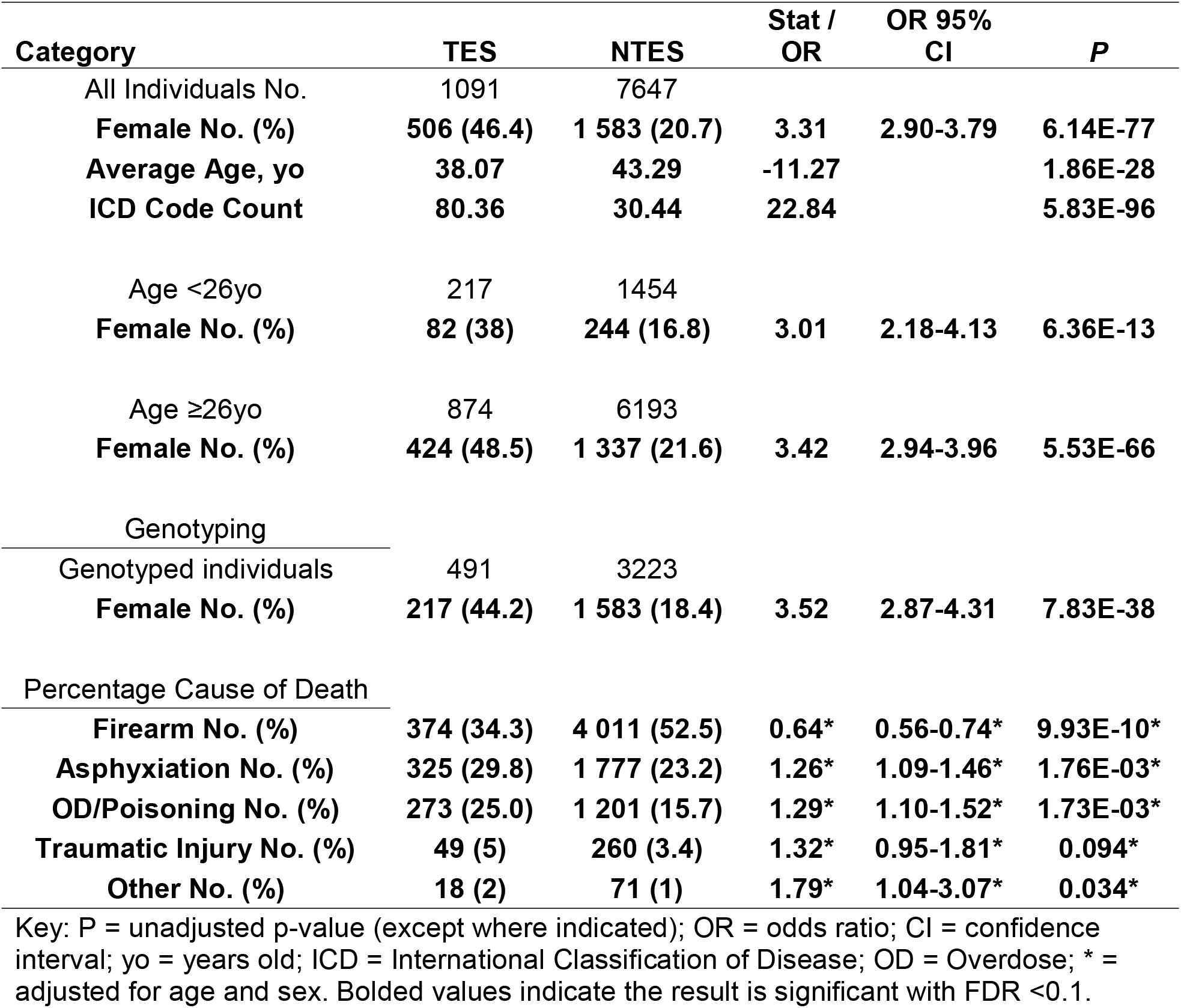
Study Population Demographics.

| Category | TES | NTES | Stat / OR | OR 95% CI | P |
| --- | --- | --- | --- | --- | --- |
| All Individuals No. | 1091 | 7647 |  |  |  |
| **Female No. (%)** | **506 (46.4)** | **1 583 (20.7)** | **3.31** | **2.90-3.79** | **6.14E-77** |
| **Average Age, yo** | **38.07** | **43.29** | **-11.27** |  | **1.86E-28** |
| **ICD Code Count** | **80.36** | **30.44** | **22.84** |  | **5.83E-96** |
| Age <26yo | 217 | 1454 |  |  |  |
| **Female No. (%)** | **82 (38)** | **244 (16.8)** | **3.01** | **2.18-4.13** | **6.36E-13** |
| Age ≥26yo | 874 | 6193 |  |  |  |
| **Female No. (%)** | **424 (48.5)** | **1 337 (21.6)** | **3.42** | **2.94-3.96** | **5.53E-66** |
| Genotyping |  |  |  |  |  |
| Genotyped individuals | 491 | 3223 |  |  |  |
| **Female No. (%)** | **217 (44.2)** | **1 583 (18.4)** | **3.52** | **2.87-4.31** | **7.83E-38** |
| Percentage Cause of Death |  |  |  |  |  |
| **Firearm No. (%)** | **374 (34.3)** | **4 011 (52.5)** | **0.64\*** | **0.56-0.74\*** | **9.93E-10\*** |
| **Asphyxiation No. (%)** | **325 (29.8)** | **1 777 (23.2)** | **1.26\*** | **1.09-1.46\*** | **1.76E-03\*** |
| **OD/Poisoning No. (%)** | **273 (25.0)** | **1 201 (15.7)** | **1.29\*** | **1.10-1.52\*** | **1.73E-03\*** |
| **Traumatic Injury No. (%)** | **49 (5)** | **260 (3.4)** | **1.32\*** | **0.95-1.81\*** | **0.094\*** |
| **Other No. (%)** | **18 (2)** | **71 (1)** | **1.79\*** | **1.04-3.07\*** | **0.034\*** |

Key: P = unadjusted p-value (except where indicated); OR = odds ratio; CI = confidence interval; yo = years old; ICD = International Classification of Disease; OD = Overdose; \* = adjusted for age and sex. Bolded values indicate the result is significant with FDR <0.1.

### Genotyping

Individual blood samples were processed and genotyped as fully described elsewhere.^10^ Briefly, whole blood was processed to extract DNA, which was then genotyped via Illumina Infinium PsychArray (https://www.illumina.com/products/by-type/microarray-kits/infinium-psycharray.html). Individuals were removed if there was excessive missing data (missing call rates of greater than 0.01), high relatedness to another subject (p-hat > 0.125), and < 90% European ancestry. Individual variants were removed if they violated Hardy-Weinberg equilibrium with p < 1.0E-06, were not successfully called in ≥ 10% of subjects, or if they had a minor allele frequency (MAF) of < 0.01. Remaining individuals were subjected to imputation via the Michigan Imputation Server,^26^ utilizing the Haplotype Reference Consortium reference panel. Following imputation, variants were removed with poor imputation quality (R^2^ < 0.5, AvgCall < 0.9) or which had a MAF < 0.01.

### Genetic Analyses

PGS were calculated from the statistically weighted genotype results of forty phenotypes from eighteen large, publicly available Genome Wide Association Study (GWAS; see Supplemental Table S4). Phenotypes were selected based on TES clinical findings. Of note, suicide attempt datasets that included USMRS data^11,27^ had this data removed prior to statistical weight and PGS calculations. PGS were calculated using the PRS-CS tool ^28^ with global shrinkage parameter (□) of 0.1, MCMC iteration count of 2 000, and burn-in of 1 000 to address small discovery samples. Output data were transformed to Z-scores. These scores were compared for correlation across all phenotypes and are represented in Supplemental Figure S1, representing substantial correlations across phenotypes. The asoecation between TES/NTES group membership and polygenic scores were assessed using logistic regression models, with TES/NTES status as the dependent variable and PRS score for the given phenotype as the independent variable. Secondary models were also generated across age groups and within male and female only individuals. All models were adjusted for sex, unless the model was in male or female-only individuals, age, and the first 10 ancestry principal components. All tests were corrected for multiple testing for 26 independent phenotypes (based on intrasample correlations for many phenotypes, see Figure S1) with an FDR < 0.10 considered significant. All statistical analyses were performed within R. ^25^

## Results

### Primary evaluation: TES versus NTES, all individuals

Population demographics are summarized in Table 1. The TES group was observed to be significantly more female (46.4% versus 20.7%; Odds Ratio (OR) = 3.3, 95% Confidence Interval (CI) =2.9-3.8), younger at time of death (38.1yo vs. 43.3yo) and have more diagnostic ICD codes (95.4 versus 40.1 ICD codes per individual) compared with the NTES group. For method of death, the TES group was significantly less likely to die from firearms (34.3% versus 52.5%; OR = 0.64, 95% CI = 0.56-0.74) and were more likely to die from asphyxiation and overdose (29.8% and 25% versus 23.2% and 15.7%, respectively; OR range = 1.26-1.29) as compared with the NTES group.

ICD-based analyses of primary clinical categories (Figure 1A; Table S5) showed that suicidal behavior, socioeconomic, substance use, behavioral health, injury, and sleep-related ICD code categories were significantly more prevalent in TES group than the NTES group (OR range = 1.3-3.7). Conversely, somatic pain, gastrointestinal, renal/genitourinary, respiratory, cardiac, developmental, metabolic, and cancer-related diagnoses were more prevalent in the NTES group (OR range = 0.59-0.85).

**Figure 1:**
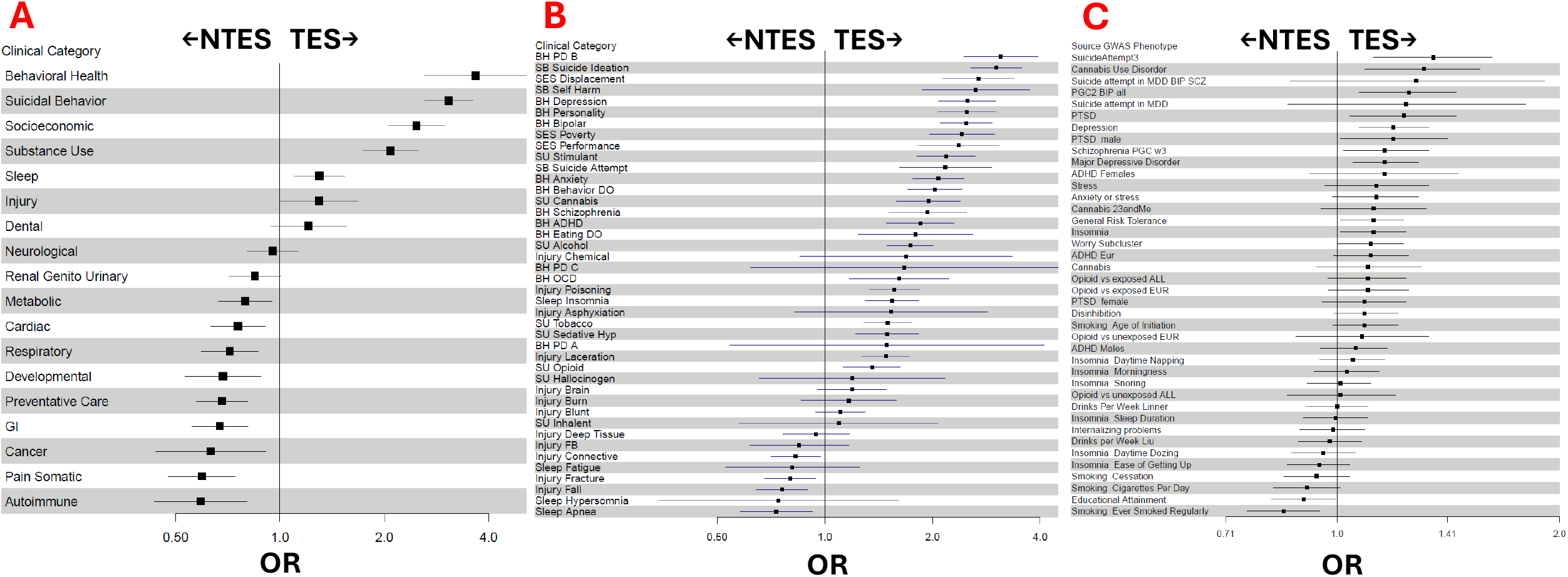
Forest plot representations of all TES versus NTES individuals. A) primary clinical categories, B) clinical subcategories, and C) genetic polygenic risk assessment. The X-axis represents odds ratio. The vertical line represents an odds ratio of 1.0, with category results appearing on the left side of the line being overrepresented within NTES individuals and category results appearing on the right side of the line being overrepresented within TES individuals.

ICD-based subgroups (Figure 1B; Table S6) enriched in TES included behavioral health diagnoses, driven by “cluster B” personality, depression, bipolar, and anxiety disorders. In addition, intentional self-harm ideation and behaviors, including suicidality, and all socioeconomic indicators were elevated in TES (OR range = 2.2-3.0). Multiple substance use diagnoses were enriched in TES (OR range = 1.4-2.2). Injury-related subcategories were largely evenly distributed among groups, but lacerations and poisoning injuries were enriched in the TES group (OR range = 1.5-1.6), and connective tissue, fracture, and fall-related injuries were enriched in the NTES group (OR range = 0.73-0.83). Insomnia prevalence was higher among TES individuals (OR = 1.5, 95% CI = 1.3-1.8) and apnea prevalence was higher among NTES individuals (OR = 0.73, 95% CI = 0.58-0.93).

Finally, genetic evaluation demonstrated an association between increasing PGS with odds of being TES derived from studies focused on depression,^29,30^ bipolar disorder,^31^ cannabis use disorder,^32^ suicide attempt,^11^ insomnia,^33^ worry/neuroticism,^29^ general risk tolerance,^34^ schizophrenia,^35^ and PTSD ^36^ (OR range = 1.1-1.4) (Figure 1C; Table S7). Only two categories, educational attainment ^37^ and smoking,^38^ showed increased polygenic scores in the NTES group (OR range = 0.85-0.90).

### Secondary Analyses: TES versus NTES with stratification by age, sex, and age/sex

Results from stratified models are summarized here, with full results being presented as follows: stratification by age is presented in Figure 2 and Supplemental Tables S8 - S11; stratification by sex is presented in Figure 3 and Supplemental Tables S12 - S15; stratification by both age and sex is presented in Supplemental Figure S2-S4 and Supplemental Tables S16 - S19.

**Figure 2:**
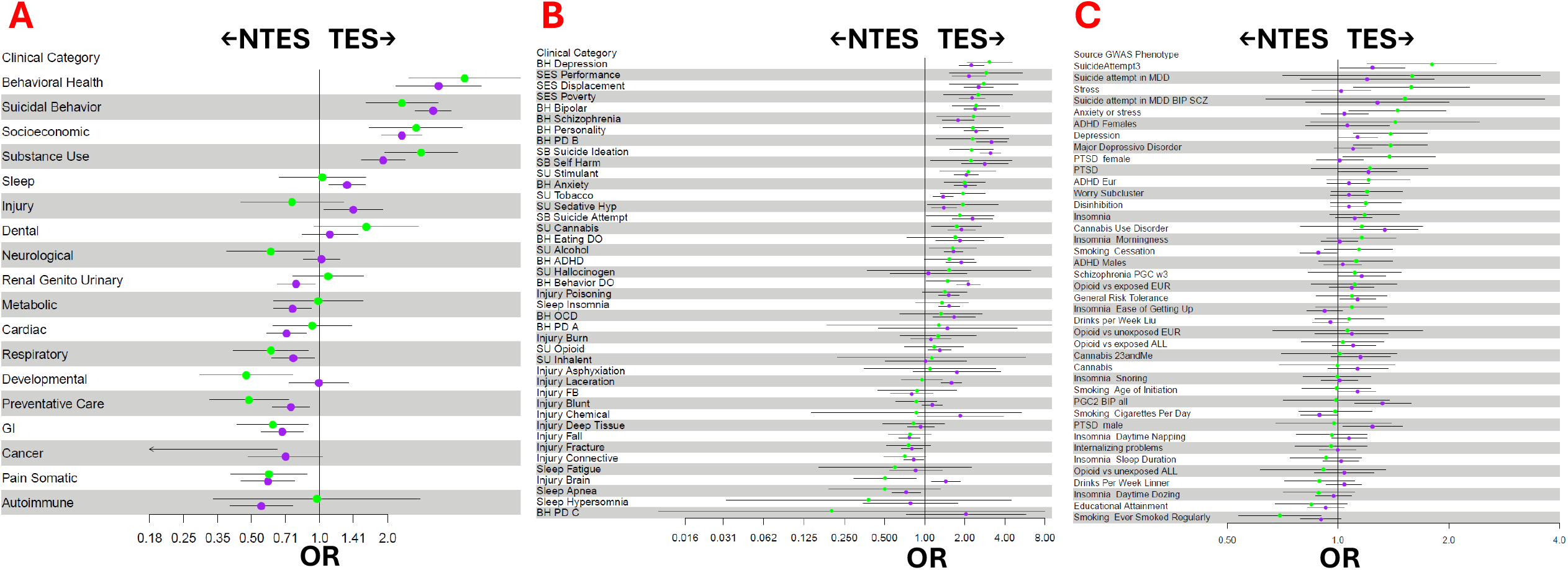
Forest plot representations of all TES versus NTES individuals, stratified by age. Green markers indicate individuals <26yo and purple markers indicate individuals ≥26yo, assessed for A) primary clinical categories, B) clinical subcategories, and C) genetic polygenic risk assessment. The X-axis represents odds ratio. The vertical line represents an odds ratio of 1.0, with category results appearing on the left side of the line being overrepresented within NTES individuals and category results appearing on the right side of the line being overrepresented within TES individuals.

**Figure 3:**
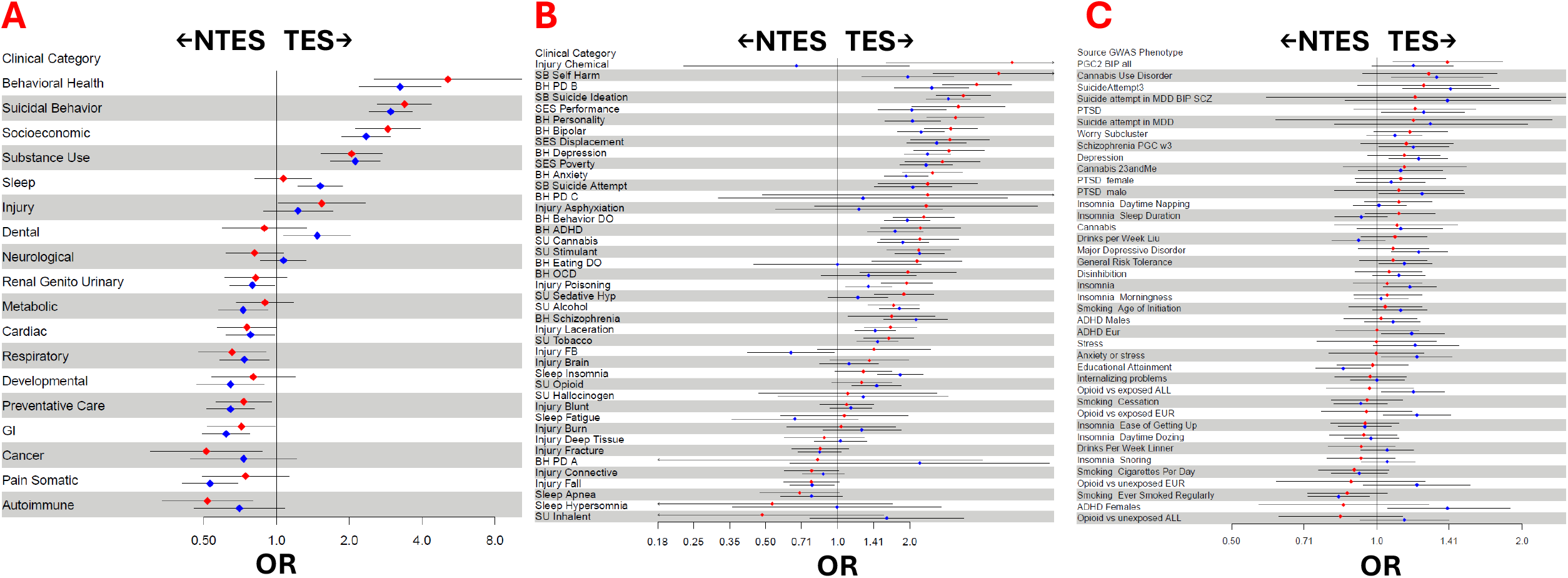
Forest plot representations of all TES versus NTES individuals, stratified by sex. Red markers indicate female individuals, and blue markers indicate male individuals, assessed for A) primary clinical categories, B) clinical subcategories, and C) genetic polygenic risk assessment. The X-axis represents odds ratio. The vertical line represents an odds ratio of 1.0, with category results appearing on the left side of the line being overrepresented within NTES individuals and category results appearing on the right side of the line being overrepresented within TES individuals.

Briefly, clear differences in method of death were observed. Use of asphyxiation was more common in <26yo and overdose within ≥26yo female individuals in both TES and NTES groups, but both methods were consistently more prevalent across TES individuals (OR_asphyxiation_ = 1.3-1.4, and OR_overdose_ = 1.5-1.9). All TES were less likely to use firearms for suicide than NTES, regardless of sex or age (OR range = 0.6-0.8). Diagnostic patterns showed uniquely increased prevalence of cluster B personality disorder in <26yo female TES versus NTES individuals (OR = 3.5, 95% CI = 2.7-5.5) and of bipolar disorder in <26yo male TES versus NTES individuals (OR = 3.3, 95% CI = 2.1 - 5.4). Sleep disorders were more prevalent in male TES versus NTES individuals (OR = 1.7, 95% CI = 0.99 – 2.8) and less prevalent in female TES versus NTES individuals (OR = 0.39, 95% CI = 0.16 – 0.91). For substance use, <26yo TES versus NTES individuals had greater prevalence of tobacco use disorder (OR = 1.94, 95% CI = 1.3-2.9), but ≥26yo TES versus NTES individuals showed particularly high prevalences of stimulants, alcohol, and cannabis (OR range = 1.7-2.1). Male TES versus NTES uniquely showed increased prevalence of opioid use disorder (OR = 1.5, 95% CI = 1.2-1.8) while female TES versus NTES uniquely had increased prevalence of sedative/hypnotic use (OR = 1.9, 95% CI = 1.4-2.5). Finally, PGS genetic signals differed between TES and NTES particularly based on age, with <26yo TES showing increased PGS in depression, anxiety/stress, and PTSD (OR range = 1.4-1.6), and ≥26yo TES showing increased PGS for bipolar and cannabis use disorders (OR range = 1.32-1.34). In addition, male TES individuals uniquely showed increased PGS for ADHD and opioid dependence (OR range = 1.2-1.4).

## Discussion

To our knowledge, this study is the first to simultaneously evaluate clinical and genetic factors associated with interpersonal trauma exposure among post-mortem suicide decedents, made possible by the unique USMRS resource. Numerous differentiating characteristics between groups appear to converge on specific comorbidities that vary by age and sex. Results offer key insights that may help direct personalized approaches that target specific risks, depending on population characteristics. In addition, the significant, and sometimes directionally opposing variation observed by age and sex, such as with disordered sleep prevalence in males and females, support the importance of utilizing stratified analyses across demographic characteristics and warrant caution with “lumped” analyses. Essentially, findings were nuanced and suggest different clinical risk factors may or may not be important depending upon the specific at-risk population subset.

Importantly, results highlight critical differences in the trajectory to suicide death within those previously exposed to interpersonal trauma and may point to valuable and implementable interventions. Because TES individuals, on average, die from suicide *several years sooner* than NTES individuals, timely and targeted interventions are especially needed for this population. TES individuals were also observed to more frequently pursue less violent and potentially interruptible means of suicide death, including asphyxiation and overdose. Therefore, strategies such as increased engagement, supervision, and/or regular check-ins could be particularly high impact prevention strategies. Specific assessment and counseling focused on medication stockpile restriction could also be important, especially among female trauma-exposed individuals, with greater risk with increasing age. In addition, TES individuals demonstrated consistently higher numbers of diagnostic codes. This pattern may indicate greater average engagement with healthcare services, and thus, there may be more abundant opportunities to intervene among TES.

Clinically, TES individuals were significantly more likely to have behavioral health diagnoses, particularly depression, bipolar disorder, and anxiety, and this pattern was most pronounced among those <26yo. Substance use disorders were also overrepresented in TES individuals, most notably tobacco use among those <26yo and stimulant, alcohol, and cannabis use among those ≥26yo. These results underscore the role of comorbid psychiatric conditions and substance misuse in potentially mediating the pathway from trauma to suicide. Alternatively, psychiatric symptoms may be misattributed to an underlying psychiatric disorder, but arise secondary to traumatic exposures, leading to poor treatment response and designation as a “treatment resistant” patient. Observed variation in substance use by age in the TES group could reflect aspects of accessibility of specific substance types, diagnostic bias, and/or specific biological interactions, such as increased sensitivity and developmental impacts for nicotine use among adolescents and young adults.^39^ Such findings reinforce the importance of screening for substance use in individuals with prior interpersonal trauma.

In the U.S., suicide mortality exhibits a persistent sex bias, with higher rates among males despite higher prevalence of suicidal ideation and attempts among females.^2^ Yet, the female suicide deaths in the TES group (46.4%) highlights a subgroup in which female suicide mortality is substantial. This underscores the need for sex-responsive prevention strategies. Sex-stratified analyses revealed differences in diagnostic patterns, with specific variations evidenced based on age. Specifically, <26yo TES females had increased diagnoses of personality disorders while <26yo TES males had increased diagnoses of bipolar disorder. Sex-specific patterns may reflect biological susceptibility differences and/or gendered sociocultural factors influencing trauma processing, care-seeking behavior, and expression of psychopathology that influences diagnostic patterns and biases. In particular, the fact that diagnoses of personality and bipolar disorders become considerably more frequent in ≥26yo TES males and females may hint at divergent trajectories. These findings align with literature indicating that females more likely report trauma and are diagnosed with PTSD, but that expression, comorbidity, and management of trauma-associated symptoms may be more nuanced across the sexes.^40-42^

Polygenic score results were completely distinct between age groups, suggesting differences in underlying biology. Specifically, TES individuals <26yo have more diagnoses and biological loading for depressive, anxiety, and trauma associated disorders. Given that younger individuals exposed to trauma tend to not respond consistently or effectively to traditional psychopharmacology approaches for depression,^43-45^ such results may indicate a need to encourage more rapid implementation of trauma informed evidence-based psychotherapies in these patients. ^46,47^

## Limitations

There are several limitations of this study to consider. First, the study relies on ICD coding which can lack sensitivity for trauma.^48^ Emerging evidence suggests that natural language processing can improve identification of key psychiatric historical elements.^49,50^ However, missed cases make results conservative due to unidentified cases within the comparison group. Future work using natural language processing or instrument data will allow more powerful and nuanced analyses, yet our results represent an important starting-point. In addition, a comparable control group with genetic data was not available for this study, limiting our ability to test relevant resilience factors. Future studies that utilize matched control populations with and without trauma exposures would be helpful in addressing such questions. Additionally, sex-stratified genetic analyses were constrained by smaller sample sizes, limiting statistical power. As a result, null findings in female-specific analyses should be interpreted cautiously. Finally, the population of Utah is largely White Non-Hispanic, and though this sample was representative and population-derived, this limits generalizability. This issue is particularly pronounced with respect to the genetic analyses, which is restricted to European individuals due to employed methods being highly sensitive to population structure. Future studies incorporating more diverse populations would allow greater generalizability.

## Conclusions

This study represents the first evaluation of clinical and genetic correlations within interpersonal trauma-exposed suicide deaths. The results provide critical and potentially actionable differences within individuals with prior interpersonal trauma who die from suicide. Additionally, evidence suggesting differing biological liability may also provide guidance for more personalized suicide prevention interventions for trauma exposed individuals. Together, these findings suggest that trauma is not merely risk factor but may define a distinct suicide phenotype. These differences may reflect unique etiological pathways that impact neurodevelopment and dysregulation versus specific genetic vulnerabilities. Perhaps most important, TES individuals die at younger ages, more often use substances, seek more care, and tend to use less lethal means for suicide death. These factors argue for early trauma detection, targeted case management, and use of trauma informed evidence-based care. Ongoing work should capitalize on improved trauma identification to develop more meaningful comparisons to define resilience factors and mitigate identified TES specific vulnerabilities to reduce suicide death.

## Data Availability

All data produced in the present study are available through formal affiliation with the Utah Suicide Mortality Research Study and the University of Utah.

## Acknowledgements

Funding for work presented in this study arose from NIH grants R01MH122412 (Coon PI) and R01MH123489 (Coon PI), the Brain and Behavior Foundation Young Investigator Grant 31248 (PI Monson) and generous support from the Bertin Family Foundation. Partial support for all datasets within the Utah Population Database was provided by the University of Utah Huntsman Cancer Institute and the Huntsman Cancer Institute Cancer Center Support grant, P30 CA2014 from the National Cancer Institute. Research was supported by the NCRR grant, “Sharing Statewide Health Data for Genetic Research” (R01 RR021746, G. Mineau, PI) with additional support from the Utah Department of Health and Human Services and the University of Utah. We thank the University of Utah Clinical and Translational Science Institute (CTSI) (funded by NIH Clinical and Translational Science Awards), UPDB staff, University of Utah Information Technology Services and Biomedical Informatics Core for establishing the Master Subject Index between the Utah Population Database, the University of Utah Health Sciences Center, and Intermountain Healthcare. We also thank the Utah Suicide Mortality Research Study (USMRS), the Utah Office of the Medical Examiner, NIH (R01MH122412) and the University of Utah’s Office of the Vice President for Research for supporting and processing data related to suicide death. We also thank Intermountain Healthcare for data access and data support. The support and resources from the Center for High Performance Computing at the University of Utah are also gratefully acknowledged. The authors wish to thank the Utah Department of Health and Human Services (DHHS) for providing data for this publication, specifically from the Healthcare Statistics Program. The authors are solely responsible for the analyses, interpretations, and conclusions presented, which do not necessarily reflect the opinions or policies of the Utah Department of Health and Human Services or the state of Utah. Finally, we thank the Utah Department of Health & Human Services and the University of Utah Office of the Vice President for Research for support of this work.

